# The Time Required for Primary Care Consultations: Estimates of the Expected Duration of Initial Sick Child Visits in Low- and Middle-income Countries Using the Integrated Management of Childhood Illness (IMCI) Clinical Algorithm

**DOI:** 10.1101/2023.06.03.23290926

**Authors:** Anthony Carter

## Abstract

**Background:** Little is known about the time required for primary care consultations, particularly those involving sick children in low- and middle-income countries (LMICs). This paper begins to fill that gap by providing evidence-based estimates of the time needed for initial visits with under-five infants and children at public or not-for-profit facilities in LMICs that use the Integrated Management of Childhood Illness (IMCI) clinical algorithm. The estimates are evidence-based and case-mix adjusted.

**Methods:** The expected duration of IMCI consultations in any given setting is a function of the health issues presented by patients less than 5 years old, the kind and number of tasks health workers must perform to care for such patients, and the time required to complete those tasks. Determining the time needed to complete tasks with no minimum duration requires, in addition to tabulations of health issues and lists of tasks to be performed, information on rates of task performance and the mean observed duration of consultations. All this information was located, first, by searching MEDLINE, the database of the International Network for Rational Use of Medicines, the websites of the WHO and its regional offices, GOOGLE, and GOOGLE SCHOLAR using search terms such as ‘Integrated Management of Childhood Illness’, ‘observational’, ‘prospective’, ‘classification’, ‘clinical signs’, ‘health facility survey’, ‘validity’, ‘algorithm’, and ‘chart booklet’. Additional information was found by reviewing work that cited a qualified study and, conversely, material included in the bibliographies of qualified studies.

**Results:** Estimates of the expected mean duration of initial consultations were constructed for 39 patient populations in 22 countries. The mean ranges from 17.1 to 20.5 minutes in the infant populations and from 15.5 to 26.4 minutes in the child populations. The shortest consultations range from 16.1 to 16.7 minutes in the infant populations and from 11.0 to 17.4 minutes in the child populations. The 98^th^ percentile of expected durations ranges from 21.5 to 25.7 minutes in the infant populations and from 22.5 to 35.2 minutes in the child populations.

**Conclusions:** IMCI consultations with infants and children presenting for the first time with a new concern require considerably more time than previously recognized. This suggests that the personnel costs of providing appropriate IMCI services may be substantial. It also confirms that the quality of most IMCI consultations is low.

## Introduction

Little is known about the time required for primary care consultations, particularly those involving sick children in low- and middle-income countries (LMICs). This paper begins to fill that gap by providing estimates of the time needed for initial visits with under-five infants and children at public and not-for-profit facilities in LMICs that use the Integrated Management of Childhood Illness (IMCI) clinical algorithm. The estimates are evidence-based and case-mix adjusted.

Health care providers have a great deal to do during primary care visits. They must begin the consultation, establish rapport, elicit the patient’s concerns, gather information about presenting illnesses, perform a physical exam, discuss diagnosis and treatment, summarize, and bring the consultation to a close [1, 2{Byrne, P, 1976}]. The time required to accomplish all this remains something of a mystery. In general, it is thought that consultations rarely should be shorter than ten minutes [3, 4]. It also is thought that longer consultations generally are superior to shorter ones. Longer visits do not guaranteed appropriate care [4], but in Europe and North America physicians in longer consultations recognize more issues, explore more psychosocial concerns, give fewer prescriptions, and/or provide more preventive measures [5–12]. Similarly, during longer consultations in Tanzania, clinical officers and assistant medical officers ask more questions and perform more examinations [13]. Longer consultations often are essential for those with multiple morbidities [3]. Finally, several experienced physicians in the United States and Argentina regard 15 to 20 minutes as the optimal duration of primary care visits [14, 15].

It has been suggested that the time required for IMCI consultations falls in the lower half of this range. A leading member of the team that designed the IMCI algorithm suggests that IMCI consultations “can be completed in 10-15 min although some cases take longer” [16]. According to the World Health Organization (WHO), the mean duration of sick-child consultations in primary care facilities where the IMCI algorithm is used is 15 minutes [17].

The problem with all of this is that it rests on little or no evidence. None of it takes account of the mix of problems with which patients present [12, 18], the tasks involved in patient care, or the time required to perform such tasks.

The IMCI clinical algorithm and the research surrounding it provide unusually rich resources with which to address these issues. A comprehensive program to reduce “unacceptably high” infant and child mortality in LMICs, IMCI was developed in the mid 1990s by the WHO and the United Nations Children’s Fund and has been widely adopted [19, 20]. IMCI seeks to encourage proper health practices in families and communities, to strengthen health care systems, and to improve the technical quality of primary care services. The IMCI clinical algorithm is the key means used to achieve the last of these aims, Concerned with health issues frequently found in resource-poor settings [16, 21, 22], the algorithm integrates a variety of interventions shown to be cost-effective in previous vertical programs [23]. For infants less than 2 months old, it focuses on bacterial infections, feeding problems, low weight, and, in some versions, jaundice. For children 2-59 months old, the foci include acute respiratory infections, especially pneumonia; diarrhea; fevers, especially malaria and measles; malnutrition, and anemia. Immunization status is a concern for both age groups. The algorithm provides a scheme to identify the health issues presented by under-5 patients, tasks to be used in the care of such patients, and criteria by which tasks are elicited. Crucially, research on the design and use of the algorithm, its effects on provider performance, and related topics furnishes the information needed to estimate the expected duration of initial consultations with sick children and infants.

## METHODS

The expected duration of IMCI consultations in any given setting is a function of the health issues presented by patients less than 5 years old, the kind and number of tasks health workers must perform to care for such patients, the time required to complete those tasks, and, in a few instances, the organization of work within facilities. An estimate of the time needed to perform tasks that have no predetermined duration requires information on health issues presented, tasks to be performed, task-specific performance rates, and the mean observed duration of consultations.

### Definitions

#### Health Issues and Classifications

Health issues are conditions presented by patients that require providers to perform tasks.

IMCI classifications are categories used to organize signs and symptoms indicative of the character, occurrence, and severity of health issues that account for a substantial portion of child mortality in low- and middle-income countries. Classifications differ from “etiologically distinct diagnoses” in two ways [24]. Some combine distinct diseases into a single category. “For example, a child with the classification ‘very severe febrile disease’ could have meningitis (viral or bacterial), … sepsis (from several possible pathogens), … cerebral malaria,” pertussis, plague, or pneumonia. Conversely, a single disease may be assigned different IMCI classifications if it presents with different levels of severity. If pneumococcal pneumonia presents with cough and fast breathing it is classified as ‘pneumonia’; however, if it presents with cough and stridor, it is classified as ‘severe pneumonia/very severe disease’. Similarly, in areas of high malaria risk, malaria presenting with fever alone is classified as ‘malaria’. When it presents with fever and a stiff neck or unconsciousness it is classified as ‘very severe febrile disease.’

Most classifications designate one or another of the health issues with which the IMCI algorithm is concerned. However, a subset of classifications indicates aspects of health issues rather than the issues themselves. Classifications for ‘persistent diarrhea’ and ‘dysentery’ indicate aspects of diarrhea and are initially counted in classifications for ‘dehydration’. Classifications for ‘measles’ or ‘dengue’ indicate aspects of fever and are initially counted in classifications for that issue.

The labels for three health issues, ‘HIV status’, ‘other problems’ and ‘missing immunizations’, involve no classifications. One issue requiring preventive care has no label; it occurs when a child ≥2 months old but <2 years old has been given the classifications ‘not very low weight’ and ‘no anemia’.

#### Classification Profiles

Classification profiles are tabulations of the IMCI classifications, other health issues, and ancillary findings present in patient populations.

#### Tasks

A task is a piece of physical work IMCI-trained providers may have to perform during a consultation. The performance of a task requires providers to use their ears, eyes, hands, and/or voices in ways that consume time. The IMCI clinical algorithm is an arrangement of tasks.

Providers also are expected to make a variety of decisions, as in ‘Classify COUGH OR DIFFICULT BREATHING’, ‘Decide the Malaria Risk: high or low’ if a child has a fever, and ‘IDENTIFY TREATMENT’. However, in this paper, these decisions do not count as tasks because they are cognitive rather than physical and are not inherently time-consuming. Providers may sometimes take time over decisions because the issues are novel or complex or because they are uncertain or confused, but, in principle, such decisions can be made instantaneously.

#### Organization of Work

This term is used in health facility survey reports to refer to the distribution of primary responsibility for the performance of a subset of routine tasks between IMCI-trained providers and other categories of clinical staff.

#### Time Required to Perform Tasks with No Minimum Duration

The time required to perform tasks with no minimum duration is the observed duration of consultations less the time spent performing tasks with pre-defined minimum durations divided by the number of tasks with no minimum duration. Where necessary, this is adjusted for the Hawthorne effect.

### Data

#### Health Issues

Data on the prevalence of health issues, IMCI classifications, and ancillary findings on clinical signs come from gold-standard findings reported in studies of the validity and impact of IMCI, evaluations of the clinical signs used in the care of infants, and health facility surveys. Gold-standard findings are based on direct examinations of patients by researchers. In one case, information on malnutrition comes from a contemporaneous household survey.

Most of the studies used here were located by searching MEDLINE, the database of the International Network for Rational Use of Medicines, the websites of the WHO and its regional offices, and GOOGLE SCHOLAR. Search terms included ‘Integrated Management of Childhood Illness’, ‘IMCI’, ‘observational’, ‘prospective’, ‘classification’, ‘clinical signs’, ‘health facility survey’, and ‘validity’. Additional studies were identified by locating and reviewing work listed by GOOGLE SCHOLAR and other resources as citing a qualified study and, conversely, work included in the bibliographies of qualified studies.

#### Tasks and Triggering Conditions

Data on IMCI tasks come from relevant versions of the IMCI clinical algorithm. These were located using GOOGLE and GOOGLE SCHOLAR. Most of the world is covered by one or another generic version. Only a relatively small number of countries have produced their own adaptations. The key links between classification profiles and IMCI algorithms are the country in which, and the years during which, the information used in a classification profile was obtained.

In the IMCI algorithm, clinical tasks and the conditions that elicit them are indicated by a subset of sentences using imperative verbs that direct providers to carry out specific activities (see TEXT S3_NOTES ON THE IDENTIFICATION OF IMCI TASKS). Nearly all these sentences occur in Chapter 2 and Parts II-III of the IMCI handbook [25] and in the sections of chart booklets concerned with assessing, classifying, and treating infants <2 months old and children 2-59 months old (see column J, WORKBOOK S1_STUDIES USED).

#### Time Required to Perform Tasks with No Minimum Duration

The information used to estimate the mean time required to perform a task with no pre-defined minimum duration, comes from a subset of the studies used to construct classification profiles plus a paper comparing direct observation and simulated client surveys of IMCI services in Benin [26].

#### Ancillary Information on Performance of Tasks by Someone other than Provider

Where it is available, this information comes from the studies that also report the prevalence of health issues. It is based on direct observation of clinical interactions.

### Analysis

The mean expected durations of initial IMCI consultations for each patient population examined were obtained by means of the following steps (for additional details, see TEXT S1_NOTES ON METHODOLOGY).

- First, construct the population’s classification profile.
- Second, list all possible combinations of classifications and other health issues included in each classification profile, together with their probabilities.
- Third, calculate the prevalence of larger and smaller sets of co-occurring health issues.
- Fourth, identify the clinical tasks provided by each version of the IMCI algorithm and their performance conditions.
- Fifth, tabulate the number and kind of tasks required by each combination of classifications and other health issues were obtain by applying the relevant task performance conditions to the list of combinations, taking into account, where it is available, ancillary findings on clinical signs and the organization of work.
- Sixth, estimate the average time needed to complete a task with no minimum duration. NB: this was done once; the estimate was used for all patient populations.
- Seventh, calculate the expected duration of each combination of classifications and other indicators of health status by applying, as appropriate, the pre-defined minimum duration or the estimated time needed to perform tasks with no minimum duration to the required tasks.
- Eighth, calculate the content of the mean consultation in terms of the following categories: mandatory tasks with no minimum duration, mandatory tasks with a pre-defined minimum duration, conditional assessments with no minimum duration, conditional assessments with a pre-defined minimum duration, and treatment and counseling tasks.
- Finally, the expected duration for each patient population is the mean duration of all combinations weighted by their probabilities. The distribution of durations within each population also was determined.

## RESULTS

### Study Selection

Forty-seven observational studies of initial primary care consultations for children less than 5 years old report gold standard IMCI classifications, clinical signs, and/or disease diagnoses. Nine of these studies had to be excluded. Two use disease categories without indicating how these map on to IMCI classifications [27, 28]. One reports the prevalence of several IMCI symptoms but not the relevant classifications [29]. Three fail to report mild classifications for two or more symptoms and/or to distinguish among severe, moderate, and mild classifications [30–32]. Two studies report on very small subsets of classifications [31, 33]. One reports classifications for persistent diarrhea and dysentery but fails to mention dehydration [34]. A study from Nigeria [35] appears to assume that no child has more than 1 gold standard classification.

### Study Characteristics

Thirty-eight studies contain relatively complete information on the incidence of health issues. The studies were carried out between 1993 and 2017. Nine were in India; 4 in Bangladesh; 3 in Kenya; 2 each in Afghanistan, South Africa, and Tanzania, and 1 each in Benin, Bolivia, Botswana, Brazil, Burkina Faso, Cambodia, China, Egypt, Ethiopia, Gambia, Ghana, Morocco, Sudan, Uganda, Vietnam, and Zimbabwe. Since the Cambodia study includes infants <2 months old as well as children 2-59 months old, the 38 studies contain data on 39 distinct patient populations (WORKBOOK S1_STUDIES USED).

Each patient population is identified by an expression consisting of the standard three-letter abbreviation of the name of the country in which it was carried out, a 2-digit number to distinguish among studies done in the same country, and either ‘I’ or ‘C’, depending on the age group observed. For example, the 1993-94 IMCI validation survey in Kenya is identified as ‘KEN_01_C’.

Twelve of the patient populations consist of infants <2 months old. Twenty-seven consist of children 2-59 months old. For infants, the number of consultations observed ranges from 34 to 1268. For children the range is from 98 to 3200.

The original generic IMCI algorithm, in use between 1995 and 2005, applies to 7 of the 12 infant populations. The 2003/2009 India adaptation applies to the remaining 5. The 1995-2005 generic version covers all the health issues seen in 14 of the child populations and most of the issues seen in BWA_01_C, KHM_01_C, and VNM_01_C. National or provincial adaptations apply to AFG_01_C, AFG_02_C, BFA_01_C, EGY_01_C, IND_01_C, MAR_01_C, and ZAF_02_C. The 2008 generic version applies to CHN_01_C. It is assumed that the 2002 KwaZulu-Natal adaptation applies to HIV status in BWA_01_C and to all classifications in ZWE_01_C. It also is assumed that the Timor-Leste adaptation applies to dengue in KHM_01_C and VNM_01_C.

In 30 of the classification profiles, all the classifications and findings came directly from the studies used. IND_01_C, IND_03_C, and IND_08_C also required using local knowledge of the relationships between disease diagnoses and IMCI classifications. BOL_01_I, GHN_01_I, IND_02_I, IND_03_I, KEN_01_I, and ZAF_01_I were constructed, in part, by applying the relevant IMCI algorithms to reported clinical signs. BOL_01_I, GHN_01_I, IND_02_I, IND_03_I, and ZAF_01_I also required using information on the need for referral. Figures for ‘severe malnutrition’ and ‘very low weight’ in TZN_01_C come from a contemporaneous household survey in the same region. (For additional details, see Text S2_NOTES ON THE CONSTRUCTION OF CLASSIFICATION PROFILES.)

Information on the observed duration of consultations and task performance rates comes from the studies used to construct the profiles for BRA_01_C, KEN_03_C, MAR_01_C, SDN_01_C, and TZN_01_C. A study comparing the results of conspicuous observation and simulated client surveys in Benin [26] also provides a measure of observer effects (see Workbook S8).

### Health Issues

The infants and children in the 39 populations examined here presented with multiple health issues. The approach used here ignores missing information, and counts ‘other problems’, where they are noted, as a single issue.

In the infant populations, the number of issues per patient ranges from a low of 0.84 in KHM_01_I to a high of 2.48 in IND_06_I with a weighted average of 1.40 (Table 1.1). The probability that an infant presents with at least 1 health issue ranges from 0.62 in KHM_01_I to 0.97 in IND_05_I and IND_06_I (Figure 1.1). The probability that a patient presents with at least 2 issues ranges from 0.19 in KHM_01_I to 0.81 in IND_06_I. In GHN_01_I and KEN_02_I, 1 in 50 patients presents with at least 3 issues. Half of the patients do so in IND_06_I.

**Table 1.1.** Health Issues in Infant Populations.

| **PATIENT**<br>**POPULATIONS** | **POSSIBLE**<br>**BACTERIAL**<br>**INFECTION** | **JAUNDICE** | **DIARRHEA** | **FEEDING**<br>**PROBLEM OR**<br>**LOW WEIGHT**<br>**FOR AGE** | **OTHER**<br>**PROBLEMS** | **MISSING**<br>**IMMUNIZATION** | **TOTAL**<br>**HEALTH**<br>**ISSUES** |
| --- | --- | --- | --- | --- | --- | --- | --- |
| **BCD 01 I (N=234)** | 0.58 | N/R | 0.07 | 0.31 | 0.16 | NA | 1.11 |
| **BOL 01 I (N=936)** | 0.25 | N/R | 0.04 | 0.79 | 0.01 | NA | 1.08 |
| **GHN 01 I (N=535)** | 0.50 | N/R | 0.04 | 0.04 | 0.40 | NA | 0.99 |
| **IND 02 I (N=1268)** | 0.48 | N/R | 0.28 | 0.75 | 0.26 | NA | 1.77 |
| **IND 04 I (N=1268)** | 0.01 | 0.21 | 0.00 | 0.08 | 0.36 | NA | 1.32 |
| **IND 05 I (-878)** | 0.30 | 0.13 | 0.05 | 0.94 | 0.23 | NA | 1.65 |
| **IND 06 I (N=419)** | 0.55 | 0.31 | 0.13 | 0.08 | 0.28 | 0.53 | 2.48 |
| **IND 07 I (N=117)** | 0.21 | 0.08 | 0.08 | 0.70 | 0.38 | NA | 1.52 |
| **IND 08 I (N=500)** | 0.35 | 0.19 | 0.08 | 0.38 | 0.15 | 0.14 | 1.08 |
| **KEN 02 I (N=897)** | 0.74 | N/R | 0.05 | 0.39 | 0.08 | NA | 1.05 |
| **KHM 01 I (N=14)** | 0.24 | N/R | 0.29 | 0.23 | 0.09 | NA | 0.84 |
| **ZAF 01 I (N=844)** | 0.32 | N/R | NA | 0.28 | 0.32 | NA | 0.81 |
| **MINIMUM** |  |  |  |  |  |  | 0.84 |
| **MAXIMUM** |  |  |  |  |  |  | 2.48 |
| **WEIGHTED AVERAGE** |  |  |  |  |  |  | 1.40 |

Source: WORKBOOK S2\_CLASSIFICATION PROFILES FOR INFANT POPULATIONS

**Figure 1.1.**
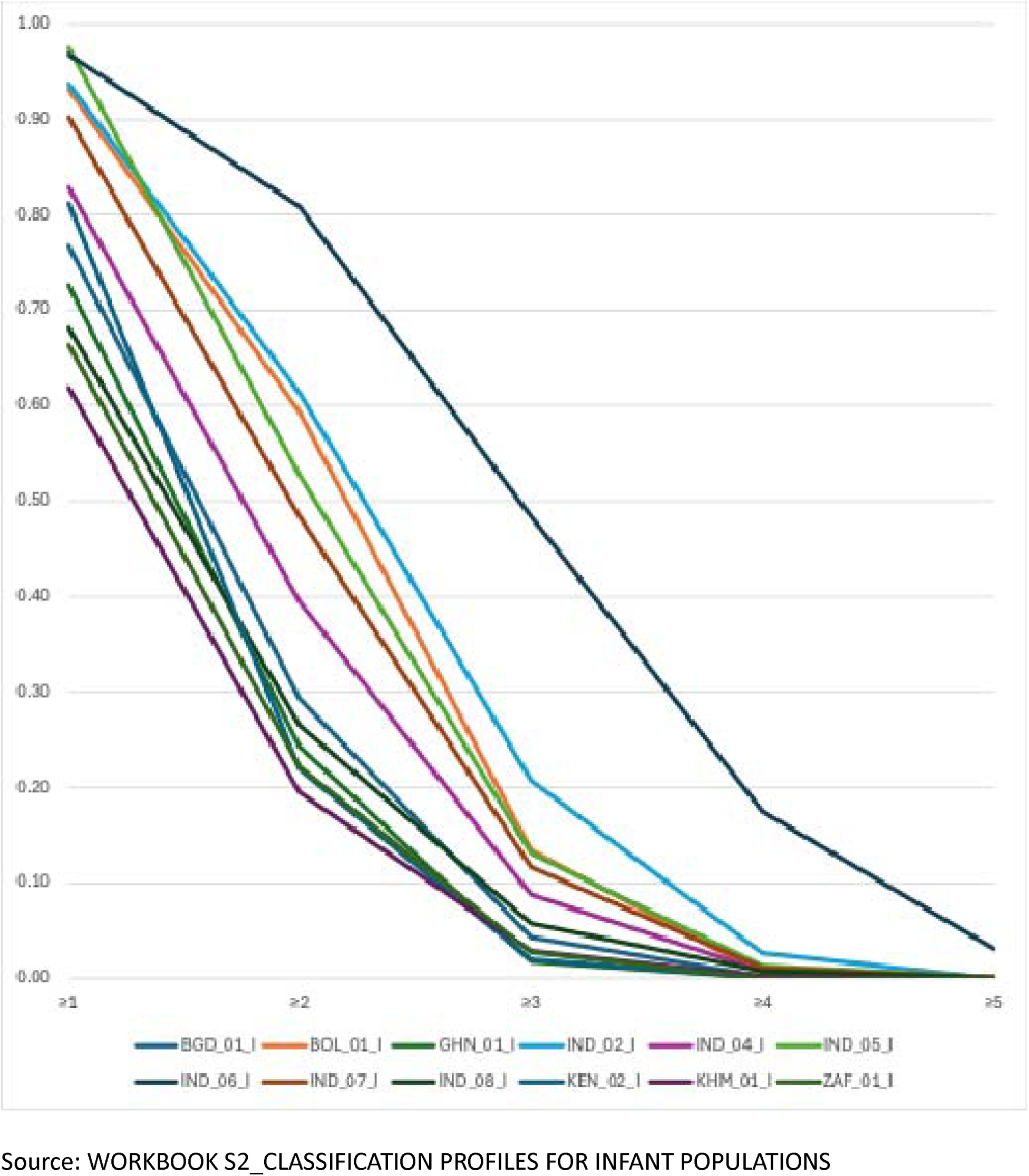
Probability of Multiple Health Issue in Infant Populations.

In the child populations, the number of health issues per patient ranges from 1.47 in IND_09_C to 3.84 in EGY_01_C with a mean of 2.44 (Table 1.2). The probability that a child presents with a least 1 issue ranges from 0.81 in IND_08_C to 1.00 in ZWE_01_C (Figure 1.2).

**Table 1.2.**
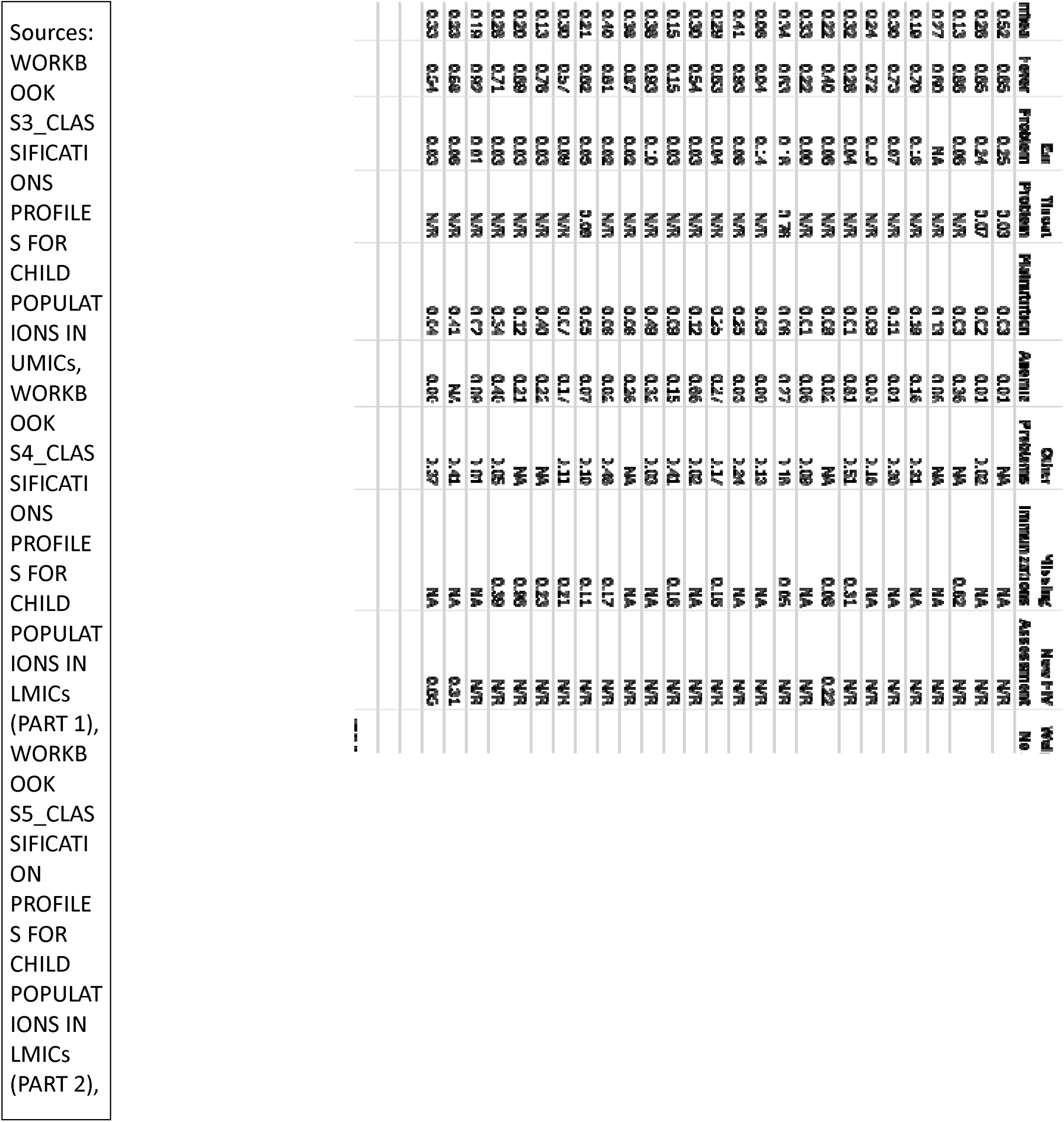
Health Issues in Child Populations.

**Figure 1.2.**
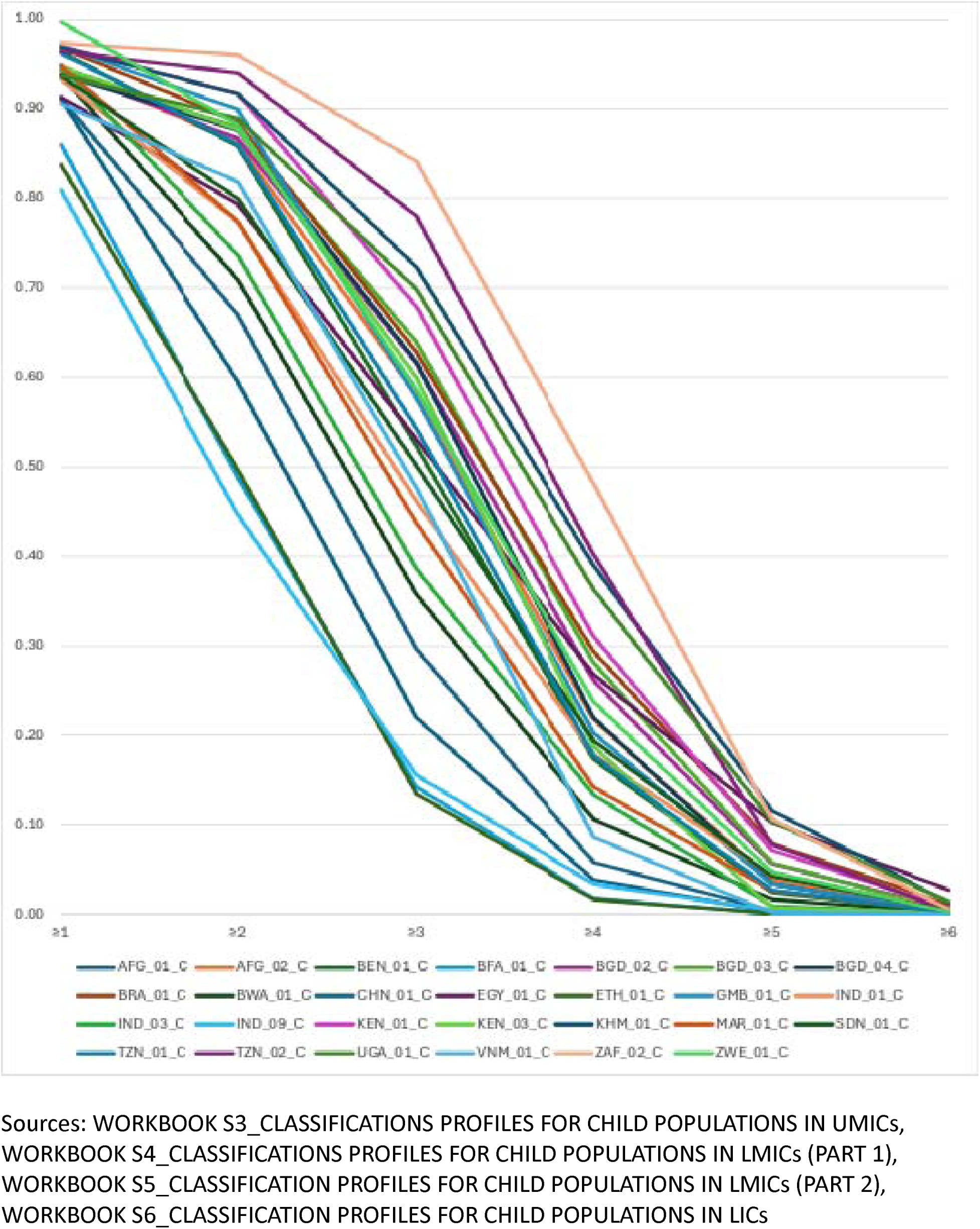
Probability of Multiple Health Issues in Child Populations.

The probability that a patient presents with at least 2 issues ranges from 0.45 in IND_08_C to 0.94 in TZN_02_C. The probability that patients present with at least 3 issues ranges from 0.14 in BFA_01_C and ETH_01_C to 0.78 in TZN_02_C. The probability that patients present with at least 5 issues ranges from 0.02 in BFA_01_C and ETH_01_C to 0.40 in TZN_02_C. In 16 of the child populations at least 1 in 50 patients present with 5 or more issues.

The kinds of issues presented vary widely from one population to another. The focus here is on issues that are used in all the populations in each age group and that are reported in all or nearly all cases. To save space, they are grouped by symptom rather than classification.

In the infant populations, the prevalence of ‘possible bacterial infection’ ranges from a low of 0.21 in IND_07_I to a high of 0.74 in KEN_02_I (Table 1.1). It is the most common issue in 6 populations. The prevalence of ‘feeding problem’ ranges from 0.04 in GHN_01_I to 0.94 in IND_05_I. It is the major issue in 5 populations. The prevalence of ‘diarrhea’ generally is lower but reaches 0.29 in KHM_01_I where classifications for ‘diarrhea’ are slightly more common than for ‘possible bacterial infection’ and ‘feeding problem’.

The most prevalent issues in the child populations are ‘cough and difficult breathing’ and ‘fever’ (Table 1.2). The prevalence of ‘cough and difficult breathing’ ranges from 0.33 in IND_08_C to 0.89 in BGD_04_C. It is the major issue in 13 populations. The prevalence of ‘fever’ ranges from 0.04 in ETH_01_C to 0.93 in GMB_01_C. It is the major issue in 12 populations. The prevalence of ‘diarrhea’ ranges from 0.06 in ETH_01_C to 0.52 in AFG_01_C, where it is the second most frequent concern. The prevalence of ‘ear problem’ ranges from 0.00 in CHN_01_C to 0.25 in AFG_01_C. The prevalence of ‘malnutrition’ is usually quite low, but is ≥0.40 in KEN_01_C, TZN_01_C, and ZAF_01_C. The prevalence of ‘anemia’ ranges from 0.00 in ZWE_01_C to 0.81 in BRA_01_C, where it is the most frequent issue.

### IMCI Tasks and their Triggers

The applicable IMCI algorithms furnish health care providers with 50 or 53 tasks for use with infants <2 months old and 95-132 tasks for use with children 2-59 months old (Tables 2.1 and 2.2). These tasks fall into three groups. Mandatory tasks are to be performed for all patients. Most conditional assessments are triggered by the results of mandatory assessments. In addition, 1 conditional assessment is triggered by the observation that a child is not awake. Another is to be performed by the provider if it has not previously been performed by intake staff. Treatment and counseling tasks are triggered by the results of mandatory or conditional assessments and in most cases are performed only when the assessment and classification process has been completed.

**Table 2.2.**
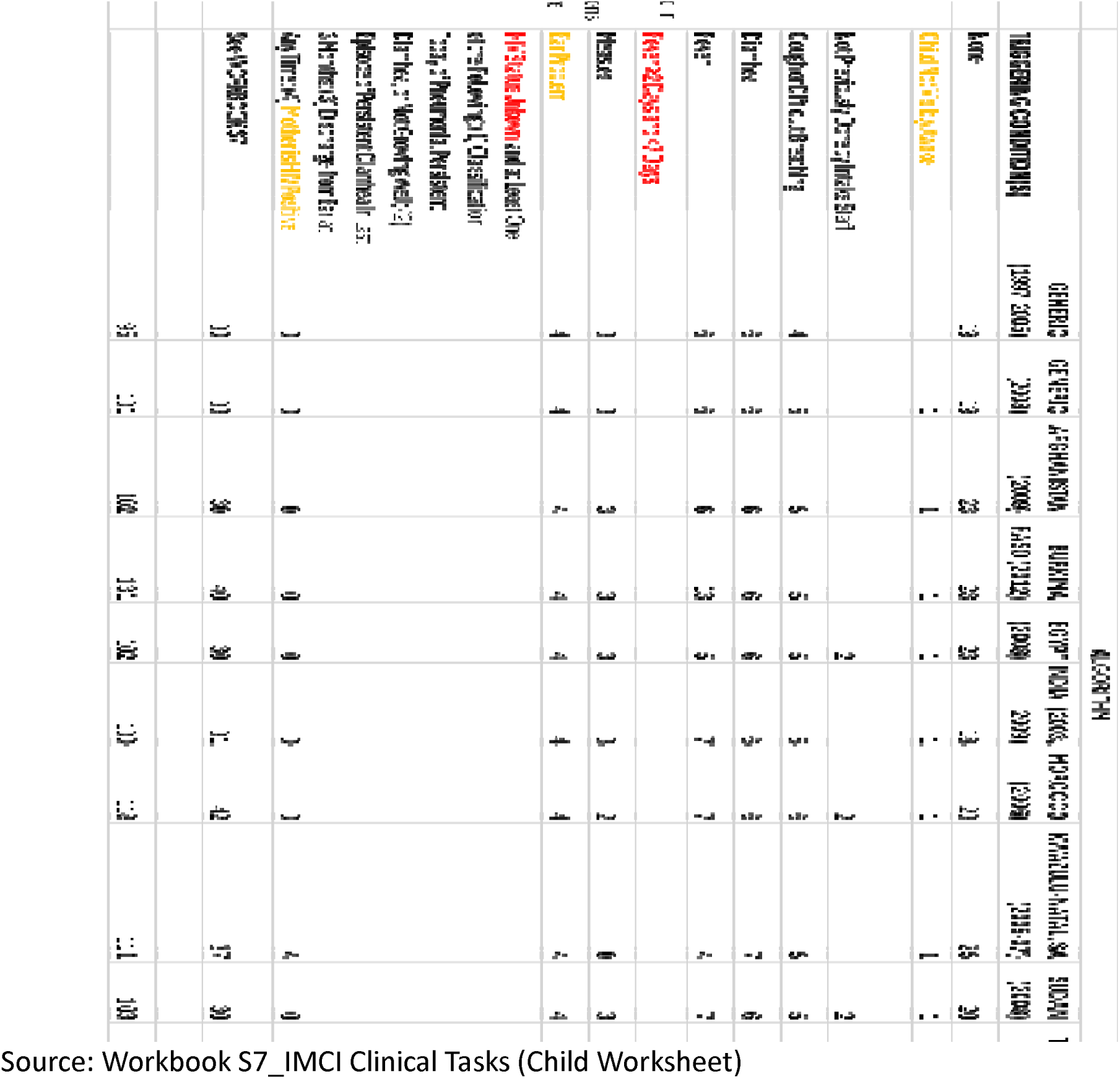
IMCI clinical tasks for children by algorithm.

**Table 2.1.** IMCI clinical tasks for infants by algorithm.

| TASK TYPE | TRIGGERING CONDITION(S) | ALGORITHM |  |
| --- | --- | --- | --- |
|  |  | GENERIC<br>(1997-2005) | INDIA (2003,<br>2009) |
| Mandatory | None | 24 | 25 |
| Conditional<br>Assessments<br>(Conditions Listed in<br>Red Are Never<br>Reported) | Initial Finding of Fast Breathing | 1 | 1 |
|  | Diarrhea | 5 | 5 |
|  | Urgent Referral to Hospital Not Required and<br>at Least One of the Following: (1) Any Difficulty<br>Feeding. (2) Breastfeeds <8 Times in 24 Hours.<br>(3) Takes Any Other Food or Drinks. (4) Low | 1 | 1 |
| Treatment and<br>Counseling | See WORKBOOK S7 | 19 | 19 |
| TOTAL |  | 50 | 53 |

Source: Workbook S7\_IMCI Clinical Tasks (Infant Worksheet)

#### Mandatory Tasks

The original generic version of the infant algorithm includes 24 mandatory tasks: a greeting and 4 opening questions, 12 assessments for possible bacterial infection, 1 check for diarrhea, 4 assessments for nutritional status, and checks for immunization status and ‘other problems’. The India adaptations provide for 25 mandatory tasks, adding 1 assessment for jaundicee, a condition treated in the generic algorithm as an ‘other problem’.

The original generic algorithm for children 2-59 months old provides 19 mandatory tasks: the 5 routine opening tasks, 3 assessments for the presence of danger signs, checks for each of the 4 ‘main symptoms’, 4 assessments for nutritional status, 1 assessment of immunization status, and a check for other problems. The number of mandatory tasks in subsequent algorithms ranges from a low of 18 in the India version to a high of 28 in the Burkina Faso version. Most of the tasks added in subsequent adaptations have to do with regional or historical differences in health issues. Throat problems are a concern in Afghanistan, Egypt, and Morocco (4 tasks). HIV is a concern in Botswana, KwaZulu-Natal, and Zimbabwe (4 tasks). Dengue fever is a concern in Cambodia and Vietnam. To shorten wait times for very sick children, KwaZulu-Natal requires providers to ‘do rapid appraisal on all waiting children’ before each new consultation.

In principle, intake staff should weigh children 2-59 months old and take their temperatures with a thermometer before they are seen by the examining health worker [25]. In most cases, however, performance of these tasks by intake staff is not reported. In these circumstances, weighing the child and checking its temperature checks are treated as mandatory tasks for the health care provider and a thermometer is not required.

#### Conditional Assessments

The original generic infant algorithms and the Indian adaptations provide 3 sets of conditional assessments. An infant found to have fast breathing should have the breaths in one minute counted a second time. A history of diarrhea triggers five assessments. Providers should perform a 4-minute assessment of breastfeeding for an infant who does not require urgent referral to hospital, but does have difficulty feeding, is breastfed less than 8 times in 24 hours, receives other foods or drinks, and/or is low weight.

The original generic algorithm for children 2-59 months old includes 23 conditional assessments: 4 for ‘cough or difficult breathing’, 6 for ‘diarrhea’, 6 for ‘fever’, 3 for ‘measles’, and 4 for ‘ear problem’. The number of conditional assessments for these issues in subsequent algorithms ranges from a low of 25 to a high of 32.

Except for the original generic version, all versions of the IMCI algorithm referenced here include a check for lethargy or unconscious, but only in children who are not visibly awake.

Health facility surveys for Egypt, Morocco, and Sudan report how many children are weighed and have their temperature taken with a thermometer by intake staff [36–38]. In these circumstances, weighings and temperature checks become conditional assessments.

KHM_01_C and VNM_01_C include classifications for dengue fever, but the applicable 1997-2005 generic algorithm provides no assessments related to this concern. Here it is assumed that the 7 conditional assessments for dengue included in the Timor-Leste algorithm are used in these settings.

The KwaZulu-Natal adaptation calls for 3 assessments to determine the HIV status of children who are not known to be HIV positive and who have at least 1 of the following: (1) a classification today of ‘pneumonia’, ‘persistent diarrhea’ or ‘not growing well’; or (2) an episode of ‘persistent diarrhea’ in last 3 months; or (3) discharge from ear at any time, or (4) ‘mother is HIV positive’.

#### Treatment and Counseling

Several of the treatment and counseling tasks provided in the relevant algorithms had to be modified before they could be used in this analysis. Three treatment options are provided for infants and children with ‘severe dehydration’. If a provider is adequately trained and if the clinic in which she works has the necessary supplies, the preferred Plan C option is to administer Ringer’s Lactate Solution or normal saline solution for 6 hours, with reassessment and adjustment every 1 or 2 hours [25]. If IV treatment cannot be provided locally or nearby, a second option is to administer ORS solution by mouth, if the infant can drink, or, if the infant is unable to drink and the provider is properly trained, by naso-gastric tube, again for 6 hours with regular reassessment. If none of these conditions can be met, the patient should be referred urgently to hospital for IV or naso-gastric rehydration. Given the constraints of training, equipment, and supplies, it is likely that the last option is the predominant first choice. In any case, it is impossible to know how, in fact, such patients are treated.

In the child algorithms that include measles, children with the classification ‘measles with eye or mouth complications’ have ‘pus draining from the eye’ and/or ‘mouth ulcers’. The algorithms provide treatments for each condition, but the observational studies report only the less specific classification. This problem is dealt with by replacing the specific treatments with ‘treat the complications of measles’.

In addition, a few treatment and counseling tasks provided for infants are triggered by feeding problems the occurrence of which is not fully reported. In this study, those tasks are replaced by ‘counsel mother about feeding issues, thrush, and/or keeping low weight infant warm’.

With these modifications, the relevant IMCI algorithms provide 19 or 22 treatment and counseling tasks for infants and 30-42 for children. Most of the differences relate to the presence or absence of health issues. The KwaZulu-Natal adaptation includes 6 tasks related HIV. The Egypt and Morocco adaptations exclude tasks related to malaria. Treatment for dengue fever may be required in KHM_01_C and VNM_01_C. Morocco also adds two counseling tasks: ‘counsel mother on how to stimulate the psychosocial development of her child’ and ‘counsel the mother on hygiene’.

### Tasks per Patient

The number of expected tasks per patient for each population is obtained by linking its classification profile and any ancillary information to the task triggers found in the applicable version of the IMCI algorithm. In the infant populations, the number of expected tasks per patient ranges from a low of 28.1 in ZAF_01_I to a high of 31.1 in IND_02_I with a mean of 29.9 (Table 3.1). In the child populations, the range is much greater, reaching from a low of 26.6 in IND_09_C to a high of 46.2 in BFA_01_C (Table 3.2). The mean is 37.2.

**Table 3.2.**
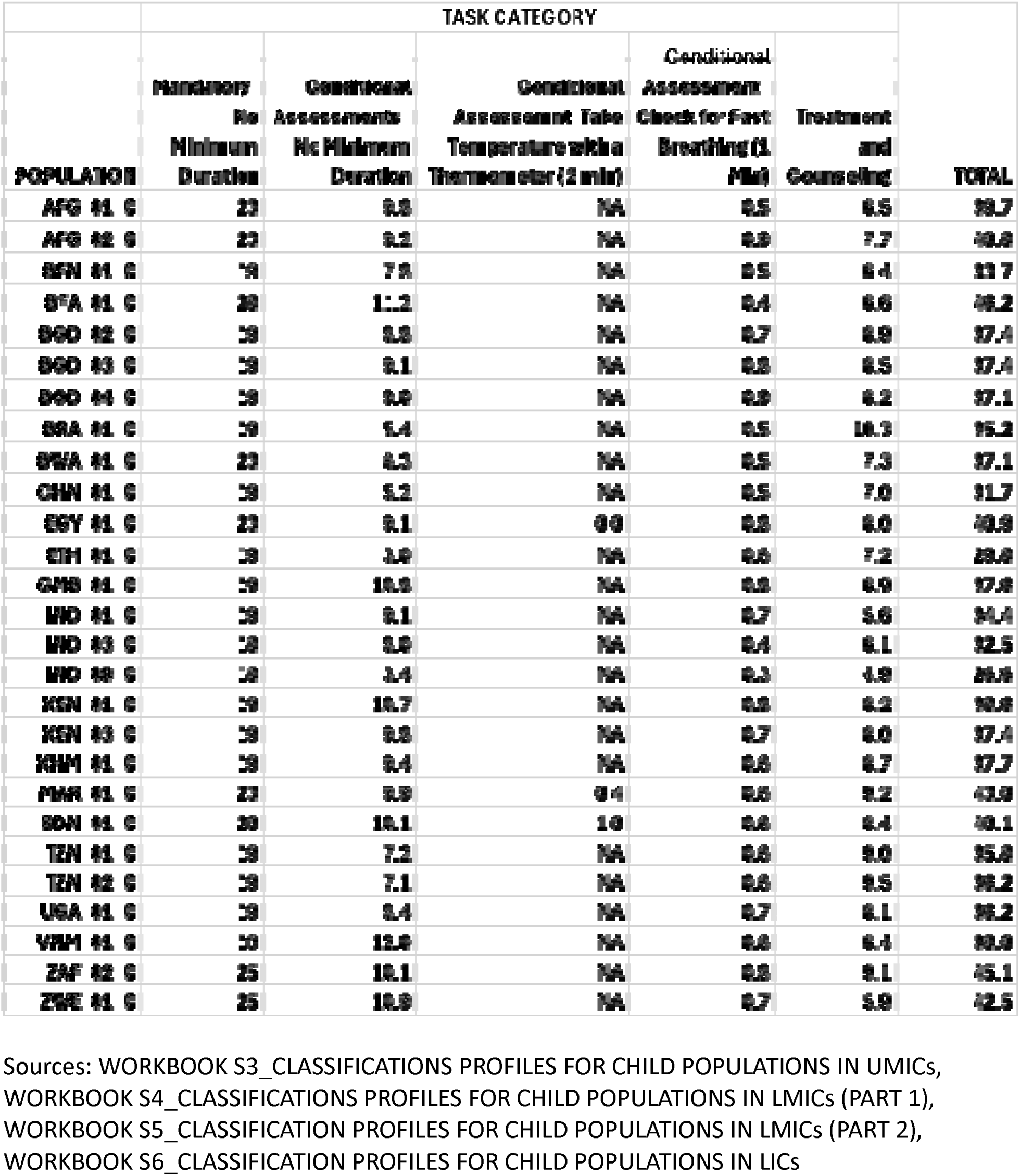
Number of expected tasks in child populations.

**Table 3.1.**
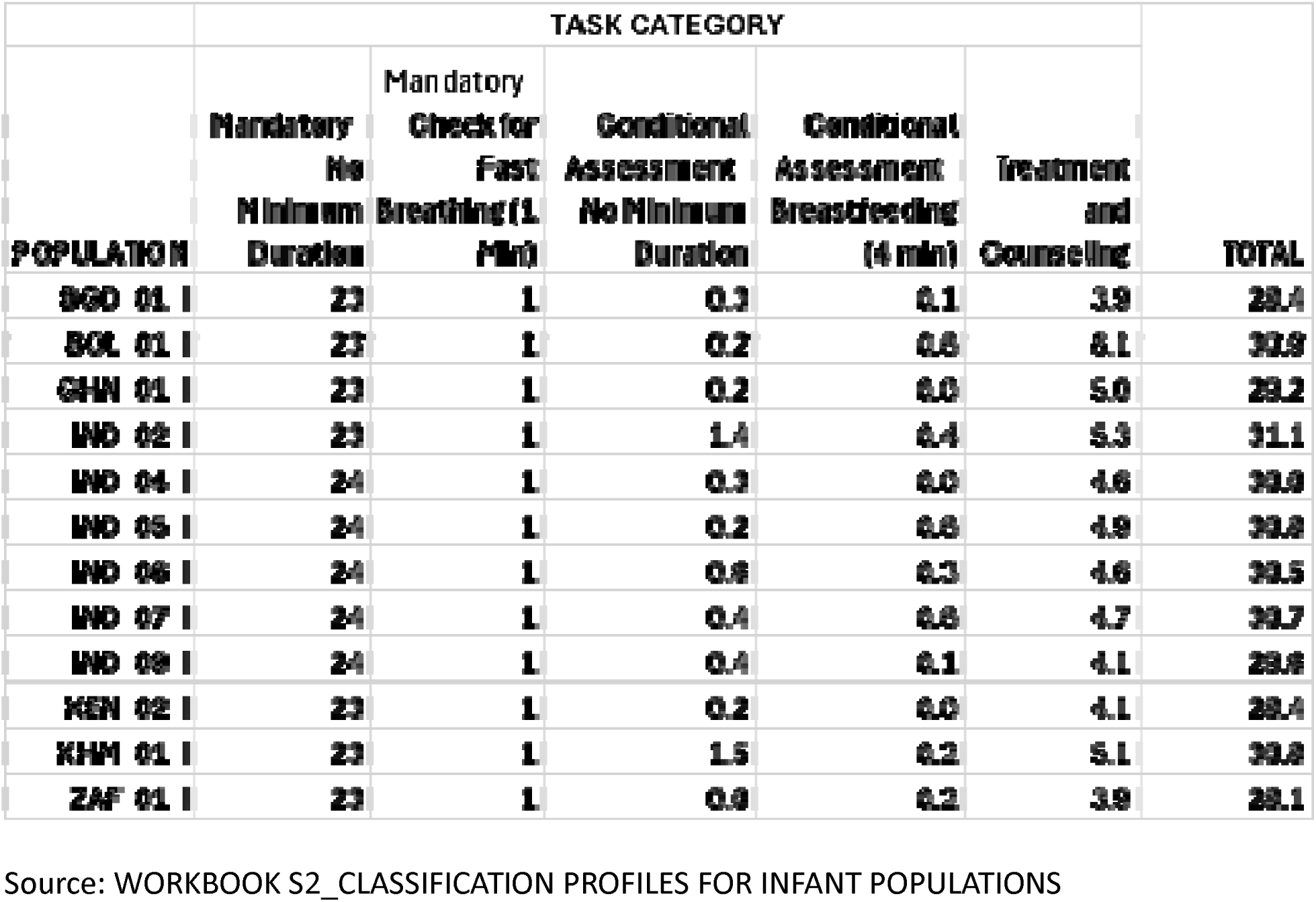
Number of expected tasks in infant populations.

### Minutes per Task

#### Tasks with Prescribed Minimum Durations

Three IMCI tasks expected have prescribed minimum durations. To assess an infant’s breastfeeding, a provider must “ask the mother to put her infant to the breast” and “observe the breastfeed for” a minimum of “4 minutes” [25]. Taking a temperature with a thermometer requires at least 2 minutes [36]. Checking for fast breathing requires at least 1 minute [25].

#### Tasks without Prescribed Minimum Durations

Table 4 provides an estimate of the time needed to perform tasks with no minimum duration. Two of the patient populations on which this estimate is based require no correction for the presence of an observer. In the Benin simulated client survey, the observer was disguised as the patient’s caretaker [26]. In BRA_01_C, providers were followed, where possible, for a 5-day working week and no difference was found in the duration of consultations between the first and subsequent days [39]. In the other cases, the minutes required to perform a task with no minimum duration are corrected for the effects of an observer on task performance rates and on the pace of task performance, i.e. minutes used per task. The coefficients used to correct for observer effects are derived from a comparison of BENIN_C0 and BENIN_SC (see columns P-R in the first worksheet in WORKBOOK S8.)

**Table 4.** Mean minutes per task with no minimum duration weighed by task occasions observed.

| **PANEL 1: MINUTES PER TASK WITH NO MINIMUM DURATION** |  |  |  |  |  |
| --- | --- | --- | --- | --- | --- |
| **PATIENT POPULATION** | **CONSULTATIONS OBSERVED** | **NUMBER OF TASKS WITH NO MINIMUM DURATION INCLUDED IN STUDY** | **PERFORMANCES OF TASKS WITH NO MINIMUM DURATION OBSERVED PER CHILD** | **TOTAL TASK OCCASIONS OBSERVED (B\*D)** | **MINUTES PER TASK WITH NO MINIMUM DURATION (CORRECTED FOR RATE AND PACE, IF NECESSARY)** |
| BENIN (CO) | 87 | 19 | 14.6 | 1274 | 1.4 |
| BENIN (SC) | 23 | 19 | 9.6 | 222 | 1.4 |
| BRA_01_C | 295 | 14 | 9.9 | 2907 | 0.5 |
| KEN_03_C | 553 | 32 | 27.5 | 15220 | 0.7 |
| MAR_01_C | 397 | 38 | 23.7 | 9400 | 0.5 |
| SDN_01_C | 282 | 28 | 16.4 | 4636 | 0.8 |
| TZN_01_C | 231 | 21 | 18.1 | 4186 | 0.3 |
| GEOMETRIC MEAN |  |  |  |  | 0.6 |
| **PANEL 2: QUALITY OF PROVIDER PERFORMANCE** |  |  |  |  |  |
| **PATIENT POPULATION** | **PERCENT OF PROVIDER CLASSIFICATIONS THAT MATCH THOSE OF SURVEYOR** | **PERCENT OF CHILDREN CORRECTLY CLASSIFIED** | **PERCENT OF CHILDREN GIVEN CORRECT SEVERE OR MODERATE CLASSIFICATIONS FOR 3 MAIN SYMPTOMS** | **PERCENT OF CHILDREN PRESCRIBED UNNECESSARY ANTIBIOTIC** |  |
| BENIN (CO) | 69.5 |  |  | NA |  |
| BENIN (SC) | 43.0 |  |  | NA |  |
| BRA_01_C | NA | 65.3 |  | 7.3 |  |
| KEN_03_C | 68.9 |  |  | NA |  |
| MAR_01_C | NA |  | 76.6 | 23.6 |  |
| SDN_01_C | 49.1 |  |  | 37.4 |  |
| TZN_01_C | NA | 63.5 |  | 14.0 |  |

Source: WORKBOOK S8 MINUTES PER TASK WITH NO MINIMUM DURATION

The mean time required by IMCI-trained providers to perform tasks with no minimum duration ranges from ranges from 0.2 minutes in TZN_01_C to 1.4 minutes in BENIN_CO and BENIN_SC. In none of these cases are providers doing satisfactory work. Performance rates for mandatory tasks, corrected for observer effects where necessary, range from 0.5 in SDN_01_C to 0.7 in TZN_01_C. Performance rates for conditional tasks are consistently worse, ranging from 0.3 in BRA_01_C to 0.5 in KEN_01_C. Substantial numbers of children are incorrectly and/or incompletely classified. In the populations for which data are available, between 7.3 and 37.4 percent of children are prescribed an unnecessary antibiotic.

Lacking a larger sample or a model case, 0.58 minutes, the geometric average of the 7 cases weighted by the number of performance occasions observed, may be taken as a reasonable first approximation of the time required to perform the average IMCI task with no specified duration. No great store should be set by this figure. However, it probably is not so high that program managers using it would be expecting too little of providers nor so low that they would be expecting providers to perform miracles of speed and efficiency.

### Expected Duration of Consultations

IMCI consultations require substantial amounts of time (Tables 5.1 and 5.2). For the infant populations, the mean expected duration ranges from 17.1 minutes in KEN_02_I to 20.5 minutes in IND_05_I. The average over all the infant populations is 18.7 minutes. For populations of children 2-59 months old, the range is from a low of 15.5 minutes in IND_09_C to a high of 26.4 minutes in BFA_01_C, with an average of 21.7.

**Table 5.2.** Expected duration of initial IMCI consultations in child populations.

| PATIENT<br>POPULATION | MANDATORY | MINIMUM | WEIGHTED<br>AVERAGE | 95TH<br>PERCENTILE |
| --- | --- | --- | --- | --- |
| IND 09 C | 10.4 | 11.0 | 15.5 | 22.5 |
| ETH 01 C | 11.0 | 12.2 | 17.2 | 23.0 |
| CHN 01 C | 11.0 | 12.2 | 18.2 | 26.0 |
| IND 03 C | 10.4 | 11.0 | 18.8 | 26.0 |
| BEN 01 C | 11.0 | 12.2 | 19.6 | 26.3 |
| BRA 01 C | 11.0 | 12.2 | 20.1 | 27.0 |
| IND 01 C | 11.0 | 12.2 | 20.2 | 28.0 |
| TZN 01 C | 11.0 | 12.2 | 20.6 | 27.0 |
| TZN 02 C | 11.0 | 12.2 | 20.8 | 29.0 |
| UGA 01 C | 11.0 | 12.2 | 21.0 | 27.0 |
| BWA 01 C | 13.3 | 14.5 | 21.3 | 29.3 |
| BGD 02 C | 11.0 | 12.2 | 21.6 | 27.6 |
| KEN 03 C | 11.0 | 12.2 | 21.6 | 27.4 |
| BGD 03 C | 11.0 | 12.2 | 21.7 | 27.1 |
| KHM 01 C | 11.0 | 12.2 | 21.7 | 27.1 |
| BGD 04 C | 11.0 | 12.2 | 21.8 | 27.3 |
| VNM 01 C | 11.0 | 12.2 | 21.9 | 29.0 |
| GHA 01 C | 11.0 | 12.2 | 22.0 | 27.6 |
| KIN 01 C | 11.0 | 12.2 | 22.4 | 27.8 |
| AFG 01 C | 13.3 | 14.5 | 23.2 | 31.3 |
| AFG 02 C | 13.3 | 14.5 | 23.7 | 31.2 |
| EGY 01 C | 13.3 | 14.0 | 24.1 | 31.2 |
| SEN 01 C | 12.2 | 15.0 | 24.3 | 32.3 |
| MAR 01 C | 14.2 | 16.3 | 24.0 | 33.2 |
| ZAF 02 C | 14.3 | 15.1 | 25.2 | 32.6 |
| ZWE 01 C | 15.1 | 16.2 | 25.6 | 33.8 |
| BFA 01 C | 16.2 | 17.4 | 26.4 | 35.2 |
| MINIMUM |  |  | 15.5 |  |
| MAXIMUM |  |  | 26.4 |  |
| WEIGHTED AVERAGE |  |  | 21.7 |  |

Sources: WORKBOOK S3\_CLASSIFICATIONS PROFILES FOR CHILD POPULATIONS IN UMICs, WORKBOOK S4\_CLASSIFICATIONS PROFILES FOR CHILD POPULATIONS IN LMICs (PART 1), WORKBOOK S5\_CLASSIFICATION PROFILES FOR CHILD POPULATIONS IN LMICs (PART 2), WORKBOOK S6\_CLASSIFICATION PROFILES FOR CHILD POPULATIONS IN LICs

**Table 5.1.**
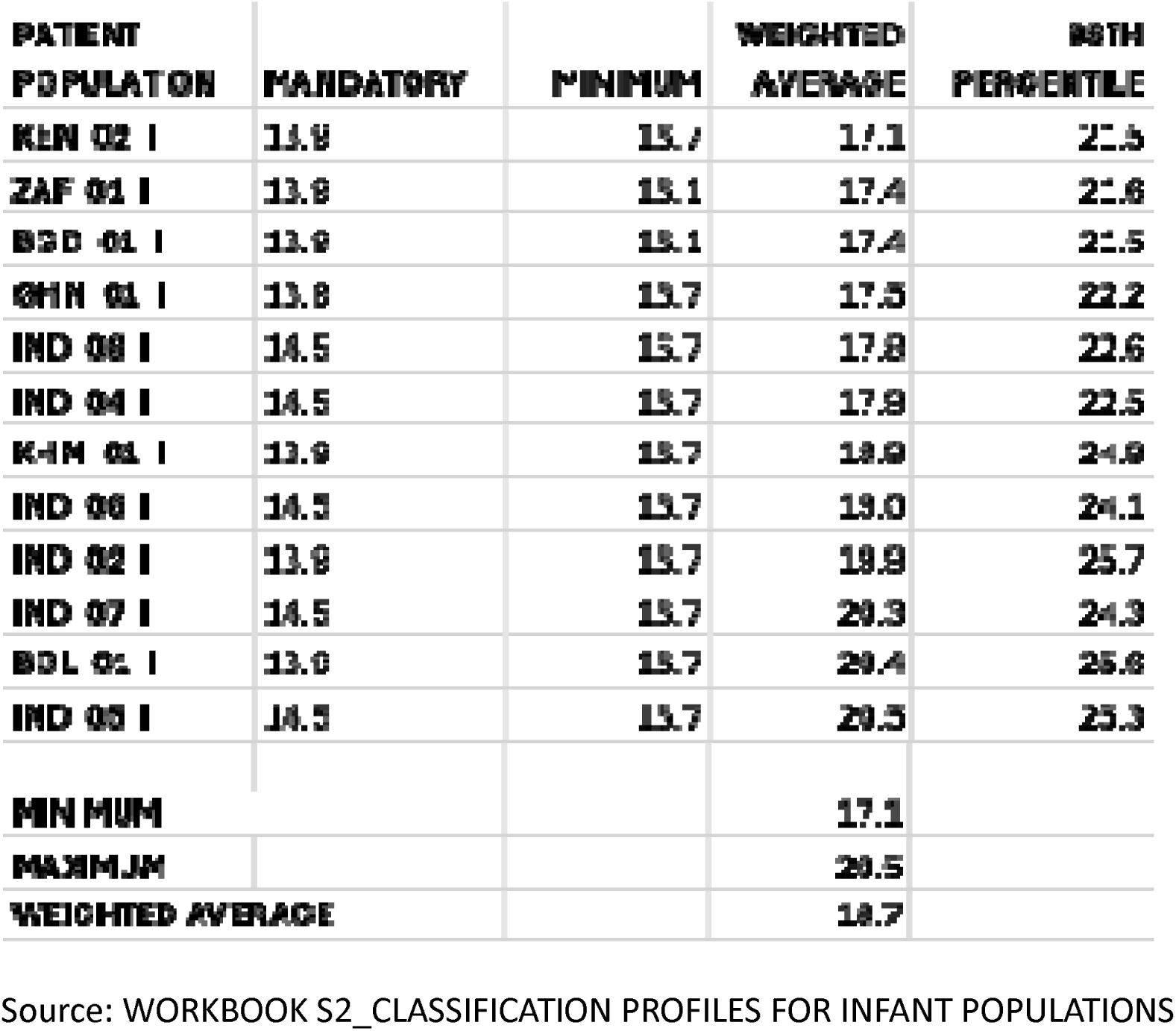
Expected duration of initial IMCI consultations in infant populations.

The shortest IMCI consultations take 16.1 or 16.7 minutes in the infant populations and 11.0 to 16.2 minutes in the child populations. This floor is determined in large part by the provision of mandatory tasks. In the original generic algorithm for infants and in the India version, these tasks are expected to take 13.9 and 14.5 minutes respectively. In the child populations, the mandatory tasks included in the India version of algorithm are expected to take 10.4 minutes. This figure rises to 11.0 in the generic versions, 12.2 in the Sudan version, 13.3 in the Afghanistan and Egypt versions, 14.2 in the Morocco version, 14.5 in the Kwa Zulu-Natal version, and 16.2 in the Burkina Faso version. The floor also includes a few treatment and counseling tasks. For example, providers are to advise all mothers when to return and to counsel them about their own health issues unless the patient requires urgent referral. Depending on the case mix presented by the patient populations and the details of the relevant algorithm, such tasks increase the minimum expected duration of consultations by 2.2 to 2.7 minutes for the infant populations and 0.6 to 2.9 minutes for the child populations.

Finally, consultations with much longer expected durations are not uncommon. In the infant populations, the 98^th^ percentile of expected durations ranges from 21.5 minutes in BGD_01_I and KEN_02_I, where it is 4.1 or 4.4 minutes longer than the mean, to 25.7 minutes in IND_02_C, where it is 5.8 minutes longer than the mean. In the child populations, the 98^th^ percentile of expected durations ranges from 22.5 minutes in IND_08_C, where it is 7.0 minutes longer than the mean, to 35.2 minutes in BFA_01_C, where it is 8.8 minutes longer than the mean. In other words, 1 in 50 consultations is likely to require substantially more time than the average.

## DISCUSSION

The expected duration of initial sick child visits in LMICs using the IMCI algorithm exceeds both our preconceptions and our experience. The PHCP anticipated that for some patients in resource-poor settings satisfactory consultations may take as little as 10 minutes [4]. However, in the infant populations examined here, the shortest expected consultations exceed this figure by 6.1 or 6.7 minutes. In the child populations the shortest expected consultations exceed the PHCP floor by 1.0 to 7.4 minutes. The WHO suggested that the mean duration of initial consultations with sick children in primary care facilities using the IMCI algorithm would be 15 minutes [17]. However, in the infant populations examined here, the mean expected duration of consultations exceeds this figure by 2.1 to 5.5 minutes. In the child populations, the mean expected duration of consultations exceeds the WHO figure by 0.5 to 11.4 minutes.

With 1 exception, the mean expected durations of initial IMCI consultations also are greater than the mean observed durations. The exception is KEN_03_C where the observed duration is 24.8 minutes (see WORKBOOK S5) and the expected duration is 21.6 minutes (Table 5.2). In this case, researchers judged the observer effect to be substantial. Providers “were clearly motivated to follow the IMCI guidelines while being observed … Thirty-seven percent of health workers acknowledged spending more time than usual with patients because of the presence of the survey team” [40]. In the other studies for which both observed and expected durations are available, the expected duration exceeds the observed duration by 9.1 minutes (81.8%) in BRA_01_C, 12.9 minutes (107.3%) in MAR_01_C, 4.3 minutes (21.6%) in SDN_01_C, and 12.4 minutes (150.9%) in TZN_01_C (WORKBOOKS S3, S5 and S6 and Table 5.2.

### Limitations

#### Missing Information

Research on IMCI services is a rich source for the investigation of the time needed to perform primary care consultations, but it is by no means perfect. A portion of the information needed to calculate the number of tasked required is missing. This information falls under 7 headings: classifications, ‘other problems’, vaccination status, age, clinical signs, the organization of work, and miscellaneous.

As regards classifications, in the infant populations information is missing for all classifications related to dehydration, persistent diarrhea, and dysentery in ZAF_01_I and for all classifications related to persistent diarrhea and dysentery in KEN_02_I. Information is missing on ‘low body temperature’ for all the populations in which it is relevant: IND_04_I, IND_05_I, IND_06_I, IND_07_I, IND_08_I, and IND_09_I. Nothing is reported for ‘local bacterial infection’ in KEN_02_I and ZAF_01_I or for ‘some dehydration’ in KEN_02_I.

In the child populations, no information is reported for any classifications related to the following: wheeze in MAR_01_C and ZAF_01_C; persistent diarrhea in BEN_01_C and TZN_01_C; dysentery in BEN_01_C; measles in BEN_01_C, TZN_02_C, and ZAF_01_C; ear problem in BFA_01_C, and anemia in ZAF_01_C. In addition, information is missing for the following individual classifications: ‘severe pneumonia’, ‘severe dehydration’ ‘severe febrile disease’, ‘severe complicated measles’, and ‘severe anemia’ in TZN_01_C, ‘mastoiditis’ in IND_08_C and TZN_01_C; ‘chronic ear infection in IND_08_C; ‘severe anemia’ in TZN_01_C; ‘symptomatic HIV unlikely’ in BWA_01_C and ZWE_01_C, and ‘≠ symptomatic HIV assessment’ in BWA_01_C.

Information on the prevalence of ‘other problems’ is reported for all infant populations but is almost certainly incomplete (for details, see TEXT S2_NOTES ON THE CONSTRUCTION OF CLASSIFICATION PROFILES). The only child populations for which information on ‘other problems’ appears to be complete and fully specified are CHN_01_C (3 conditions), ETH_01_C (7), GMB_01_C (8), IND_01_C (16), and IND_03_C (10). No information on the prevalence of ‘other problems’ is available for 10 of the 27 child populations. The figures for the remaining 12 populations appear to be incomplete.

Information on vaccination status is missing for 10 of the 12 infant populations and for 14 of the 27 child populations.

The incidence of children <2 years old is missing for 10 of the 27 child populations.

As regards clinical signs in infant populations, if a first, mandatory check of fasting breathing finds ≥60 breaths in 1 minutes, a second, conditional check for fast breathing is required. In the populations using the India infant algorithm, finding that a mother has pain while breastfeeding triggers checks for flat, inverted or sore nipples and for engorged breasts or breast abscesses. Neither of these findings is reported.

In the child populations, information is missing on the following clinical signs.

- In the Morocco algorithm as many as 7 conditional assessments are triggered by ‘wheeze’, ‘fast breathing’, and ‘chest indrawing’, but the prevalence of these signs is not reported.
- If a child has a very high fever, the provider should give paracetamol. However, the prevalence of very high fever is reported only for EGY_01_C, MAR_01_C, and SDN_01_C.
- If a child has fever lasting more than 7 days, the provider should ask if it has been present every day. Where dengue is a concern, a fever lasting ≥2 days but <7 triggers 7 conditional assessments for that disease. However, the duration of fever is never reported. It is assumed that children with a classification for dengue had a fever lasting ≥2 days but <7. Nothing is known about the duration of fever in children without such classifications.
- In all the populations except SDN_01_C and ZDN_01_C, providers who find ‘ear discharge’ should ask long it has lasted, but the prevalence of ‘ear discharge’ is not reported.
- In Burkina Faso, a child is to be offered RUTF to assess feeding if it is ≥6 months old, it has no severe classifications, and its WHF/L z-score is <-3 or it has a MUAC <115 mm. A child with the same classifications and the same nutritional findings but who is <6 months should have an assessment of breastfeeding. Neither the nutritional findings nor the ages are reported.
- The triggers for an assessment of HIV status include ‘HIV status unknown’, ‘episode of diarrhea in last 3 months’, ‘discharge from ear at any time’, and ‘mother is HIV positive’. The first 2 of these items are never reported for the populations in which HIV is a concern. The third is reported only for ZAF_01_C.
- The presence of a feeding problem triggers the task ‘counsel the mother on feeding according to the “food box” on the “counsel the mother” chart’. The prevalence of feeding problems is reported only for EGY_01_C, MAR_01_C, and SDN_01_C.
- A prolonged cough is among the signs that trigger the task ‘refer to hospital/specialist for assessment’. However, the duration of a cough is never reported.
- Signs of shock trigger 2 tasks for patients with dengue fever, but such signs are never reported.

The organization of work refers to how tasks are shared by different categories of clinic staff. The only studies that examine this topic are the health facility surveys carried out in Egypt, Morocco, and Sudan under the auspices of the WHO’s Eastern Mediterranean Region. All provide information on who weighs the child and takes her temperature with a thermometer. MAR_01_C also reports ‘check weight against growth chart’, ‘check breastfeeding status’, ‘assess use of other foods and fluids’ and ‘assess feeding practices during this illness’. EGY_02_C reports on assessments of feeding practices of children less than 2 years old without very low weight, ‘counsel the mother on feeding according to the “food box”’, and ‘advise mother when to return immediately’. It also reports on how different kinds of staff share the counseling component of ‘give course of an appropriate oral antibiotic’ and ‘rehydrate – Plan A’.

Finally, the following miscellaneous information is missing.

- In all child algorithms except for the generic original, a child who is not visibly awake is to be checked for the danger sign ‘lethargy’. However, the triggering condition is reported only for EGY_01_C, MAR_01_C, and SDN_01_C.
- In South Africa, a child with a fever should have a rapid malaria test if the test is available. The availability of such tests is not reported.
- The performance of a handful of tasks is conditional on the child being a certain age, e.g. <6 months old, but such ages are never reported.
- The tasks ‘give vitamin A’ and ‘give mebendazole’ are conditional on no dose having been given in the last N months, but information on previous treatments is never available.
- The Afghanistan, Burkina Faso, Egypt, and Morocco versions of the IMCI algorithm require an oral antibiotic to be given to children ≥2 years old and with ‘some dehydration’ if and only if there is cholera in the area. However, the presence of cholera is never reported.
- The Morocco algorithm provides ‘assess for TB’ in a child has ‘cough or difficult breathing’ or a fever and has been exposed to TB. TB exposures are not reported.
- The KwaZulu-Natal and Timor-Leste algorithms provides ‘treat cause of fever if found’ for children with the classification ‘fever – other cause’. The Timor-Leste algorithm also provides for treatment of other causes of fever, if found, in children with ‘fever-DHF unlikely’. Such causes are not reported.
- In the generic algorithms and in the Afghanistan, India, KwaZulu-Natal, and Sudan adaptations, the procedures for classifying fever vary with the risk for malaria. Malaria risk is infrequently reported.

#### Co-Morbidities

Though the synergistic relationships among nutrition, immunity, and infection are well established [21, 41] and though co-morbidities are a focal concern of IMCI [16], only 1 of the studies on which this paper is based reports patterns of association among IMCI classifications and it excludes classifications for malnutrition and anemia [42]. As a result, this study assumes that classifications and other findings co-occur randomly. This may well not be the case.

#### The Time Required to Perform Tasks with no Minimum Duration

Estimates of the time required to perform tasks with no minimum duration use information available for just 7 populations. The coefficients used to adjust these estimates for the observer effect are derived from a comparison of conspicuous observation and simulated client surveys in Benin [26]. The samples in these surveys are very small.

#### Consequences

Missing information regarding the prevalence of classifications and other findings tends to reduce estimates of the number of tasks required and thus the expected mean duration of consultations. Missing information regarding the organization of work has the opposite effect. The effects of the second and third weaknesses are unknown.

### Implications

#### Personnel Costs

The creators of the IMCI algorithm recognized that it was likely to increase the duration of consultations and, therefore, that primary care facilities might need to employ more health care providers. However, it was hoped that the increases would be modest and that they might be offset by reductions in providers’ unused capacity [16, 43].

Studies in Bangladesh [44], Brazil [39] and Tanzania [45,46] address this issue. The Bangladesh study was carried out in Matlab, the same rural subdistrict as the 3 Bangladesh IMCI evaluations used here and just a year before BGD_02_C, the first in the series. The Brazil and Tanzania studies are the sources of the data used for BRA_01_C and TZN_01_C.

The Brazil and Tanzania studies claim to provide support for the hope that adopting the IMCI program would not require hiring additional health care providers. In Brazil, observed consultations with IMCI providers initially appeared to be 71.5 percent longer than those with non-IMCI providers, 11.08 versus 6.46 minutes. However, after controlling for other determinants of consultation length such as time of day, IMCI visits were just 29.2 percent longer [39]. In Tanzania, the average consultation with an IMCI-trained provider was 30.2 percent longer than those in comparison facilities, 8.2 minutes as opposed to 6.3 [45].

As we have seen, however, observed consultations generally are significantly shorter than expected consultations. This is a consequence of the fact that observed task performance rates are low. In Brazil, where the results do not have to be adjusted for the presence of an observer, IMCI-trained providers performed, on average, 84 percent of 10 mandatory tasks and just 48 percent of 5 conditional tasks [39]. In Tanzania, where the results do have to be adjusted for observer effects, IMCI-trained providers performed, on average, 91 percent of 7 mandatory tasks and 72 percent of 6 conditional tasks [45]. After adjustment, these figures fall to 69 percent for mandatory tasks and 45 percent for conditional tasks. As we have seen, the mean expected duration of consultations is 21.2 minutes in BRA_01_C and 21.3 minutes in TZN_01_C (Table 5.2).

In the Bangladesh study, visits with paramedics and community health workers using a questionnaire based on the IMCI protocol were, on average, well over twice as long as those in which providers used their pre-IMCI practices, 18.8 minutes versus roughly 8 minutes. As the authors note, this suggests that adoption of the IMCI clinical algorithm is likely to result in a “significant increase” in personnel costs [44].

The estimates of the mean expected duration of initial IMCI consultations offered here provide strong support for the Bangladesh study. The estimates for BGD_02_C, BGD_03_C, and BGD_04_C exceed that for the original Bangladesh study by 2.8 to 3.0 minutes (Table 5.2). Just 2 of the 27 estimates for populations 2-59 months old fall below 18.8 minutes: IND_09_C, by 2.7 minutes and ETH_01_C, by just 1.6 minutes (Table 5.2).

In short, the personnel costs associated with adopting the IMCI algorithm likely are substantial. In the absence of adjustments for task performance rates, the observed duration of consultations should be understood not as an indicator of the time required for an initial sick child visit but, rather, as a crude measure of provider effort [47]. Observed consultations performed by IMCI-trained providers are longer than those performed by providers without such training but fall far short of the expected duration. Patients get the short end of the stick.

#### Time and the quality of care

Estimates of the expected duration of IMCI consultations confirm, yet again, that the quality of primary care for under-five infants and children in resource-poor settings is unsatisfactory [30, 48].

It is well established that first visits with IMCI-trained providers are “brief” [30]. A review of Service Provision Assessments (SPAs) in Haiti, Kenya, Malawi, Namibia, Nepal, Rwanda, Senegal, Tanzania, and Uganda found that, excluding “each provider’s first observed consultation to minimize the Hawthorne effect” as well as “the longest 1 percent of visits”, the duration of visits ranged from 6 minutes in Malawi, Nepal, Rwanda, and Uganda to 12 minutes in Tanzania. Across the SPA studies, “82 percent of visits were shorter than 15 minutes” [30]. It should be noted, however, that SPA reports do not distinguish between initial and follow-up visits. Nor do they provide gold standard patient classifications [49,50]. As a result, though SPA surveys provide adequate information on what tasks were performed, they have little to say about what tasks were required. Nevertheless, the SPA survey results are consistent with a recent study of rural IMCI services in Cambodia, Guatemala, Kenya, and Zambia. This study found that the mean observed duration of consultations ranged from 8.2 minutes in Zambia to 12.6 minutes in Guatemala [51].

Measured against the estimates of the expected duration of IMCI consultations presented here, visits in these populations are, in fact, brutally brief. The bulk of consultations do not last long enough for health workers to perform even the minimum of mandatory tasks, let alone the full range of conditional assessments, treatments, and counseling messages.

## Data Availability

All data produced are available online at DOI: 10.5281/zenodo.13915575

## DECLARATIONS

### Ethics approval and consent to participate

Neither was required. This study did not involve human subjects, human specimens/tissue, animals of any kind, vertebrate embryos/tissues, or field research. The information used in the study was obtained from published papers and or public online sources (e.g. WHO Health Facility Survey reports). All the studies were approved by the relevant Institutional Review Boards. No additional clinical studies, surveys, interviews, or field research were conducted.

### Consent for publication

Not required. All figures and tables are original.

### Availability of data and materials

All supporting information is in the following files available at DOI: 10.5281/zenodo.13915575. WORKBOOK S1 provides references for all the studies used. DATA S1 and DATA S2 contain pdfs of health facility surveys found on the internet that have since been taken down. DATA S3 the 2015 IMCI Chart Booklet produced by the Ministry of Health in Burkina Faso copies of which were given to me by researchers.

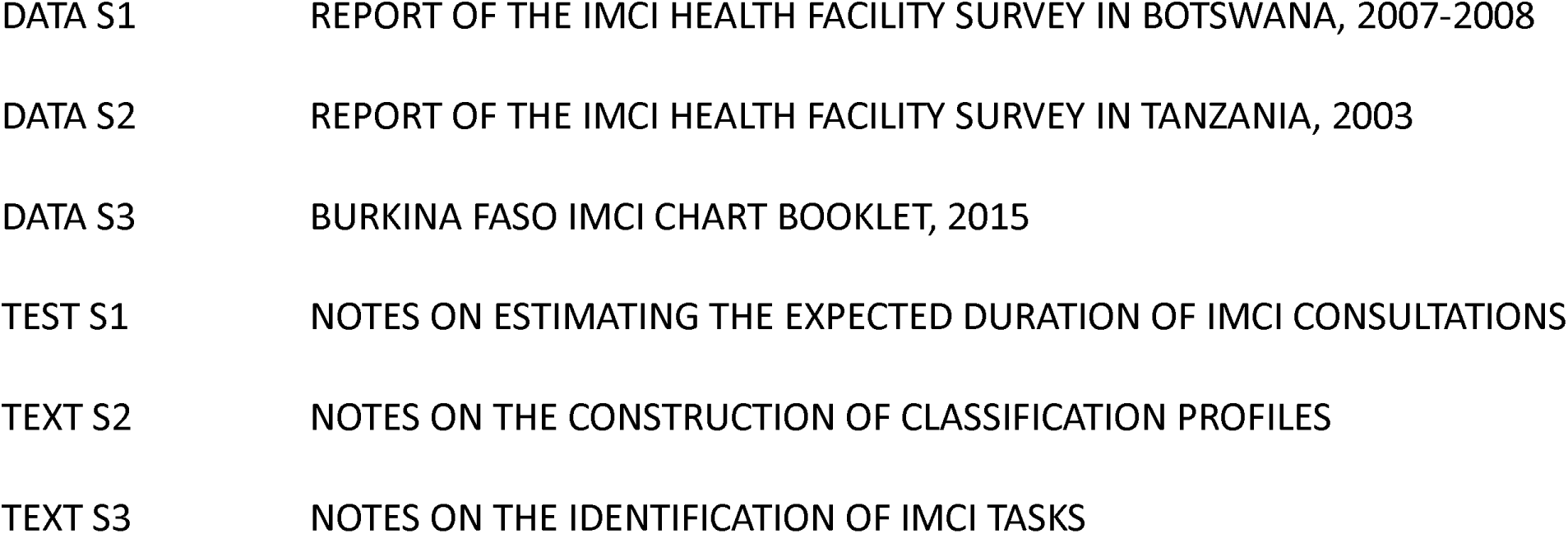
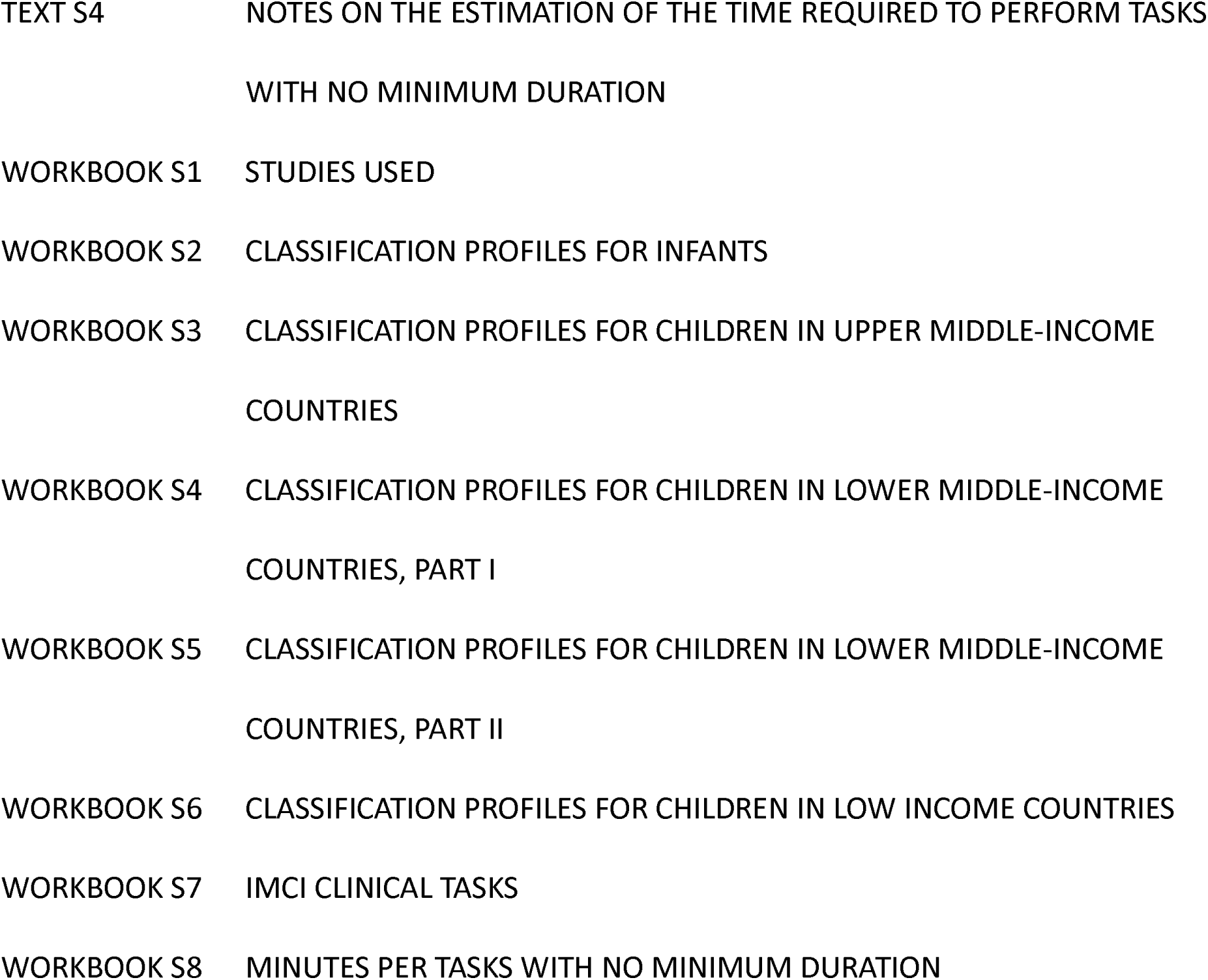

### Authors’ contributions

AC is responsible for all the work that went into this paper. He designed the study, collected and analyzed the data, wrote the paper, and wrote any required revisions.

